# Dynamic causal modeling of the COVID-19 pandemic in northern Italy predicts possible scenarios for the second wave

**DOI:** 10.1101/2020.08.20.20178798

**Authors:** Daniela Gandolfi, Giuseppe Pagnoni, Tommaso Filippini, Alessia Goffi, Marco Vinceti, Egidio D’Angelo, Jonathan Mapelli

## Abstract

The COVID-19 pandemic has sparked an intense debate about the factors underlying the dynamics of the outbreak. Mitigating virus spread could benefit from reliable predictive models that inform effective social and healthcare strategies. Crucially, the predictive validity of these models depends upon incorporating behavioral and social responses to infection that underwrite ongoing social and healthcare strategies. Formally, the problem at hand is not unlike the one faced in neuroscience when modelling brain dynamics in terms of the activity of a neural network: the recent COVID-19 pandemic develops in epicenters (e.g. cities or regions) and diffuses through transmission channels (e.g., population fluxes). Indeed, the analytic framework known as “Dynamic Causal Modeling” (DCM) has recently been applied to the COVID-19 pandemic, shedding new light on the mechanisms and latent factors driving its evolution. The DCM approach rests on a time-series generative model that provides — through Bayesian model inversion and inference — estimates of the factors underlying the progression of the pandemic. We have applied DCM to data from northern Italian regions, which were the first areas in Europe to contend with the COVID-19 outbreak. We used official data on the number of daily confirmed cases, recovered cases, deaths and performed tests. The model — parameterized using data from the first months of the pandemic phase — was able to accurately predict its subsequent evolution (including social mobility, as assessed through GPS monitoring, and seroprevalence, as assessed through serologic testing) and revealed the potential factors underlying regional heterogeneity. Importantly, the model predicts that a second wave could arise due to a loss of effective immunity after about 7 months. This second wave was predicted to be substantially worse if outbreaks are not promptly isolated and contained. In short, dynamic causal modelling appears to be a reliable tool to shape and predict the spread of the COVID-19, and to identify the containment and control strategies that could efficiently counteract its second wave, until effective vaccines become available.

## 1 Introduction

The recent COVID-19 pandemic has engendered a key debate about the factors underlying the outbreak, with the aim of finding efficient solutions to limit coronavirus dispersion. In this setting, the fight against the virus would benefit from reliable predictive models that could explain the wealth of epidemiological data in terms of a limited number of latent causes that could inform social and healthcare strategies. Italy was the first country severely hit by the outbreak outside China, particularly in its Northern part, and — similarly to, and perhaps more than other countries — has promptly adopted a tight lockdown strategy and other specific public health measures, which were successful in curbing the outbreak [Vinceti el al 2020, Bonaccorsi et al 2020]. Notably, northern Italian regions were more affected than others, with variable severity raising several questions that remain so far unanswered: i) How did the tight lockdown impact the local dynamics of SARS-CoV2 spread? ii) Why were there substantial differences across regions, despite the similar public health measures adopted? iii) Will there be a second wave? If yes, how and when should we should expect it? iv) In case of a second wave, will a further lockdown be necessary, or will efficient testing and other confinement strategies be sufficient?

Meanwhile, relevant epidemiological data have accumulated that can now be used to model future scenarios concerning the impact of COVID-19 on human lives and stress on the healthcare system [Di Domenico et al 2020, Kucharski et al 2020, Metcalf et 2020], and to identify appropriate prevention measures. In particular, epidemiological models are employed in the study of a variety of infectious diseases. Compartmental models, such as SIR-based (Susceptible – Infected – Recovered) or SEIR-based (Susceptible – Exposed – Infectious – Recovered), are commonly used to estimate transmission dynamics, the number of unreported cases and the efficacy of interventions, and have been used during the SARS-CoV-2 outbreak [Van Kleef et al 2013, Chowell et al 2016, Paiva et al 2020, Hauser et al 2020, Karnakov et al 2020, Maugeri et al 2020]. The extension of SIR models with network linkages, in which the dependencies between contacts are part of model structure demonstrates a notable improvement in the estimates of transmission parameters [Koopman 2004]. These models are generally based on differential equations modelling the rate of transition between states and effectively describe the pandemic as a dynamic system.

Recently, Friston and colleagues have proposed the application of Dynamic Causal Modelling (DCM) [Friston et al 2003] to the COVID-19 pandemic, allowing for novel interpretative perspectives on the latent factors driving the pandemic through different countries and phases [Friston et al 2020 a,b,c,d]. DCM is a flexible statistical procedure, originally designed to infer the nature of connectivity in brain networks. While its origins are in the field of neuroimaging, DCM is a generic theoretical-computational framework, which can be applied to a variety of non-linear dynamical systems, where different causal sources interact in complex ways [Friston et al. 2003]. In detail, the DCM approach involves positing an architecture of coupled causes that interact in generating observable quantities. The causes themselves are not directly observable (they are ‘hidden’ or latent), but are probabilistically inferred by the model, so to explain the observable data in a Bayes optimal fashion (i.e., as simply and accurately as possible). This methodology has the advantage of modelling the temporal trajectory of several quantities related to the process under study and simultaneously allowing a probabilistic estimation of the impact of the underlying latent factors on observable data. In DCM, compartmental models are implemented as generative models, where transitions among compartments are equivalent to pathways of information transfer within the connectome. DCM, therefore, can be aptly used to study the behavior of a network of compartments, allowing their dynamical interaction, along with the estimation of a large set of hidden parameters, under standard frameworks of Bayesian inference and parameter optimization.

The DCM approach has recently been applied to model the COVID-19 pandemic in different countries on a worldwide scale, demonstrating remarkable performance in terms of goodness of fit and predictive validity [Friston et al 2020 a,b,c,d]. Notably, DCM predicted the ongoing upsurge of cases in several European countries such as Spain, France and Germany, preluding a second wave [Friston et al 2020d]. In the present study, we adopted a similar procedure—on a finer-grain scale—by adapting it to northern Italian regions, which are characterized by a high degree of heterogeneity [Amato et al 2017]. Arguably, a DCM analysis of COVID-19 pandemic in these regions—which have been among the earlier and most strongly affected in Europe—is of special interest. We used the official epidemiological data available for each Italian region to inform model parameter estimates, while an independent data set was reserved for model validation. Using this model, in addition to characterizing the latent causes of regional differences, we aimed at predicting different scenarios for the possible second wave of COVID-19 in northern Italian regions, evincing their dependency on the efficacy of testing and tracing of infected subjects.

The present study addresses the following main aims: (1) establishing the validity of DCM in modelling the spread of COVID-19 in northern Italy, (2) inferring the unknown causal factors influencing the evolution of the pandemic in the Northern Italian regions, (3) predicting the evolution of the pandemic according to factors such as testing and tracing policies, (4) providing a reliable predictive tool that can be used to evaluate strategic interventions and implement public health measures.

## 2 Materials and Methods

### 2.1 Data sources

#### 2.1.1 Daily pandemic figures

The data reporting the pandemic evolution were obtained from the official repository of the “National Civil Protection Agency” (https://github.com/pcm-dpc/COVID-19). Specifically, we used the daily number of confirmed positive cases, deaths and recovered cases from January 22^nd^ to August 09^th^. This dataset was split into two subsets (dataset 1: from January 22^nd^ to July 20^th^; dataset 2: from January 22^nd^ to August 09^th^) to accommodate differences in the social and movement patterns before and after the end of July, a period when the great majority of the Italian population leaves home for the summer vacations.

#### 2.1.2 Cellphone-based estimates of daily movements

We also used data on daily individual movements collected anonymously via mobile cellphone networks, in order to assess the ability of the DCM to infer the probability of leaving home [Vinceti et al 2020]. Based on the position of cellphones in the six investigated regions — as available through the Call Detail Records (CDR) of SIM cards data of around 27 million Italian residents — we computed the regional daily number of cellphone movements, weighted by the provincial population. Only changes in cellphone position greater than 2 Km were considered for the computation of the movement estimates.

#### 2.1.3 Serological tests

In order to assess the validity of serological model estimates, we used the data recently made available by the Italian Ministry of Health^1^ on the seroprevalence of SARS-CoV-2 antibody positive individuals in Italy, as identified through a serological population-based survey performed by the “Italian Croce Rossa” from May 25th to July 15th.

### 2.2 The dynamic causal model

#### 2.2.1 Basic structure of the model and its applications

The DCM methodology [Friston et al 2003] was recently applied to the worldwide COVID-19 diffusion based on an epidemiological compartmental model—readers are referred to the original technical reports for details [Friston et al 2020 a,b,c,d]. In summary, the requisite generative model rests upon a mean field approximation to population dynamics, where individuals can be probabilistically characterized by their state within 4 factors, representing respectively their *location, infection, symptoms*, and *testing* state (LIST model, Fig. 1). These factors are coupled through probabilistic transitions specified by state probability transition matrices. There are 26 model parameters in total, which parameterize the initial probability of occupying each state and transitions among states. These parameters are initialized with *a priori* expectations and variances (see Table SM-1). The DCM model inversion maps from the observed data to the estimated parameter values using gradient ascent until the marginal likelihood of the data (a.k.a., model evidence) is maximized. Technically, the gradient ascent is on a variational bound on model evidence, known as evidence lower bound (ELBO) in machine learning, and variational free energy in physics. The posterior densities of the parameters returned by the procedure can be used to simulate the impact of various parameters on the dynamics of the process.

**Figure 1:**
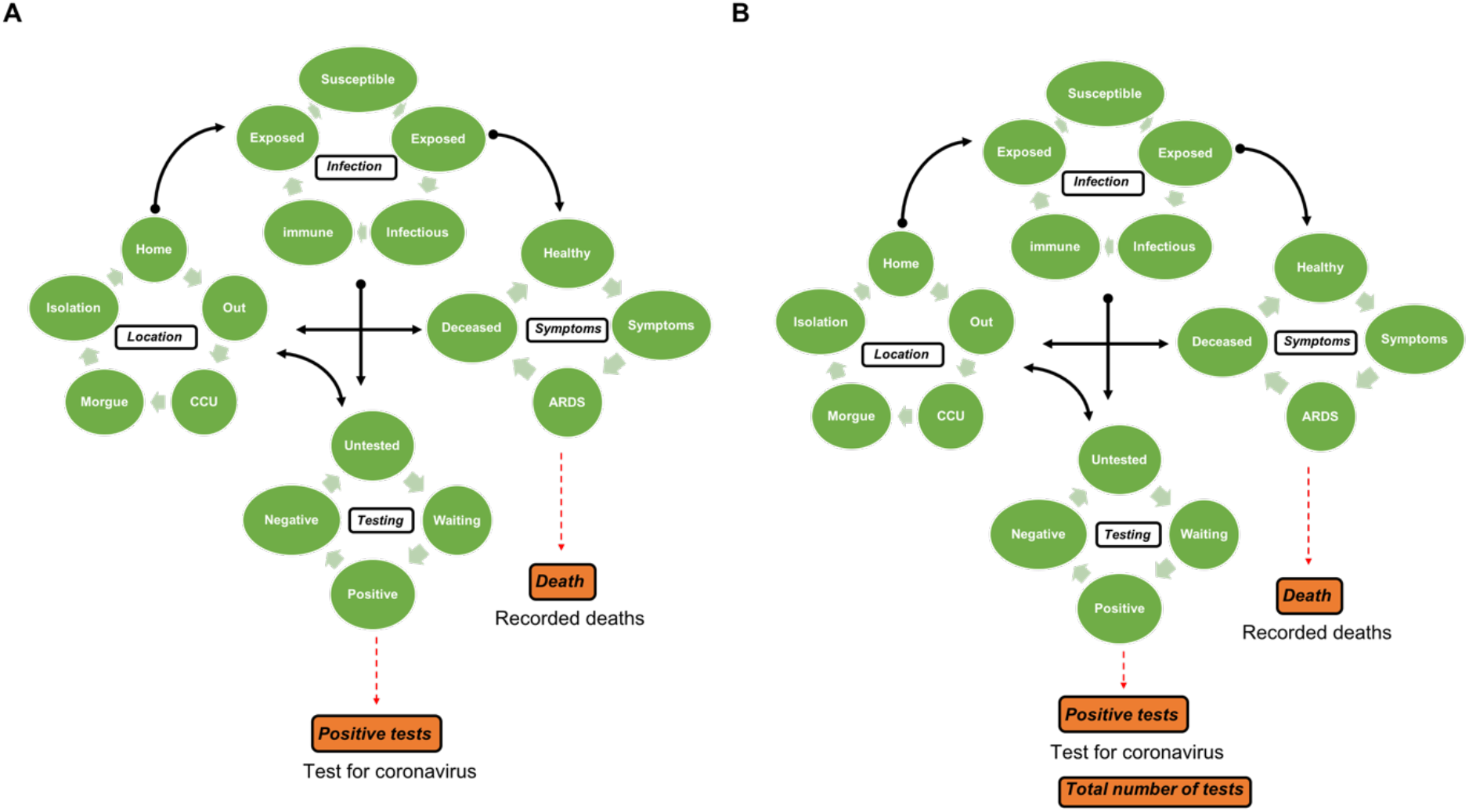
Model. The DCM model employed in this study comprises 4 distinct factors (Location, Infection, Symptoms and Testing), divided into states that characterize each individual in a population. The states within any factor are mutually exclusive, thus every individual must be in one and only one of the states associated with the each of the four factors. The transition probabilities from one state to another depend on the model parameters, which are initially specified with prior densities and are then updated through model inversion to yield posterior estimates. In the following, we provide a brief description of each factor. **Location**. Each individual can be located i) at *home* (a low contact risk location), ii) at *work* (a high contact risk location), iii) in a *Critical Care Unit* (CCU), iv) in *isolation* or v) in the *morgue*. **Infection**. Each individual can be i) *susceptible* to being infected by virus; ii) *exposed* to the virus (the individual is in contact with infected subjects) iii) *infectious* (the individual has contracted the virus and can infect other subjects), iv) *immune* (the infected individual has acquired immunity), or v) *resistant* (this is a category of people that are shielded, by virtue of host or geographical factors, from the infection). **Symptoms**. Individuals can be i) *healthy*, ii) *symptomatic*, iii) exhibiting ARDS (acute respiratory distress syndrome), or iv) *deceased*. **Testing**. Individuals can be i) *untested*, ii) *waiting* for the test outcome, iii) *positive*, or iv) *negative* on PCR testing. The factors (white rectangles) are divided into segments (states – green disks), while the orange boxes represent the observable outputs generated by the dynamic causal models (in A, daily reports of positive tests and deaths; in B, daily reports of the number of tests performed, of positive cases and deaths. In particular, the model in A is parametrized with the *effective* population size, which represents people actually in contact with contagious individuals, while the model in B is parametrized with the total (census) population size (which includes the effective population size). *(Figures adapted from* Friston et al 2020 c, d).

We adopted the same approach to model the pandemic evolution in the six Italian regions showing the maximum number of deaths during the first three months of the outbreak, namely, Lombardy, Veneto, Emilia Romagna, Liguria, Piedmont, and Tuscany. More specifically, we used DCM to the following aims:

##### Verifying the predictive ability of the model

Initially, we assessed the model’s forecast against the actual data. In order to do this, we obtained the posterior estimates of model parameters (Fig. 2A) based upon a subset of the available data, namely, the number of daily positive cases, deaths and recovered cases from January 22^nd^ to June 30^th^ and compared the model forecast to the actual data for the time points from June 30^th^ to July 20^th^. This temporal window was chosen so to exclude, for the purpose of model validation, the last week of July. Traditionally, the great majority of Italian people leave home for vacation at the end of July or the beginning of August, marking a period with significant changes in social interactions and movement patterns that could not be predicted by the model.

**Figure 2:**
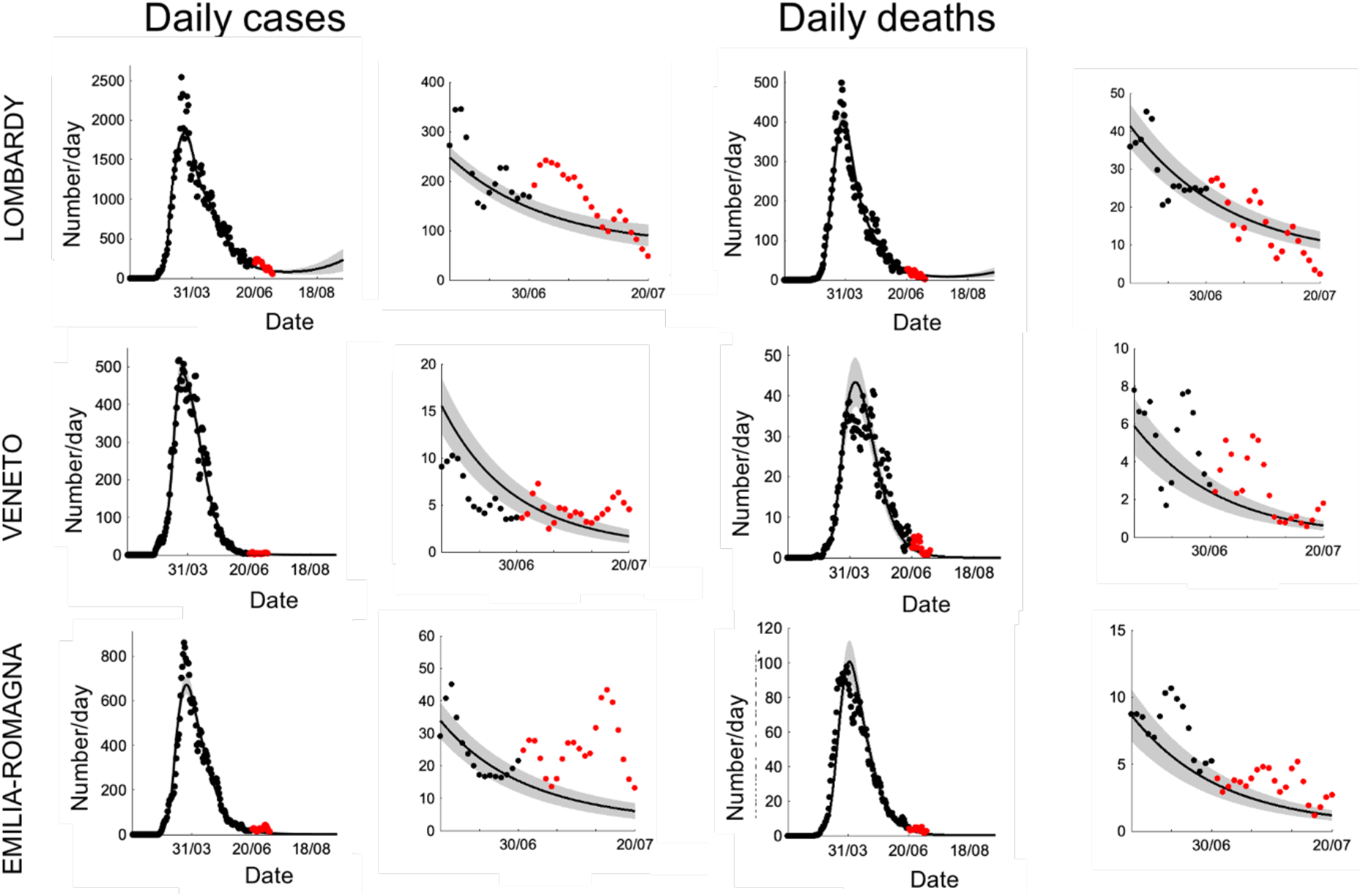
Predictive validity of the model. (Lombardy, Veneto and Emilia-Romagna). *Left*. Black dots represent daily data of positive cases reported by the “National Civil Protection Agency” for the six regions under investigation. Black lines represent the posterior expectations following model inversion. Gray bands represent the 90% Bayesian confidence intervals. Red dots correspond to data of reported positive cases from June 30th to July 20th that were excluded from data fitting, in order to validate the model prediction on the time interval shown in the inset. *Right*. Similarly, plots show the reported daily deaths from January 22nd to June 30th (black dots) and the corresponding fitting with 90% confidence interval (black lines and gray bands). Red dots represent daily deaths in the subsequent 20 days along with model prediction and 90% interval (inset).

##### Estimating the duration of immunity and its impact on the expected onsets of second waves

For this purpose, we used Bayesian Model Comparison, as described in Friston et al. 2020c. A total of 32 DCM models were specified with data comprising the number of daily positive cases and deaths (Fig. 1A) on the time interval from January 22^nd^ to August 09^th^ (dataset 2, see Methods). Each model differed in the prior assumption about the duration of immunity from 1 to 32 months in monthly increments. The model evidence for each of the 32 models was pooled over the six Italian regions under consideration, yielding a marginal likelihood of each period of potential immunity loss (PIL). It is important to note that, as explained in Friston et al 2020d, the period of immunity is not a hard threshold denoting a sudden loss of immunity. Rather, it represents instead the time constant of an exponential waning of immunity, which can depend on several factors such as population fluxes that can change the size of the susceptible pool, or factors depending on virus and hosts (see Friston et al 2020d for a detailed explanation). Since the PIL is a crucial factor in determining outbreak recurrences, its posterior estimate was used to forecast the onsets of second waves in the various Italian regions.

##### Testing and tracking strategies

This procedure (Friston et al 2020b) was used to model different aspects of testing and surveillance. The posterior estimates of model parameters (Fig. 1B) are based upon dataset 2 on daily positive cases, death and performed tests in all the six Italian regions under consideration. Crucially, the total number of tests allows a more informed parameter estimation (see Fig. SM-1).

##### Evaluating the effect of varying degrees of efficacy of the testing and tracking strategy on second-wave outbreaks

The efficacy of a testing and tracking policy is defined as the probability that a subject will be offered a test if infected and asymptomatic (Friston et al 2020b). Efficacy varies from 0 — where the probability of being offered a test when infected and asymptomatic is null — to 100% if the subject will certainly be tested and subsequently self-isolate. We performed a series of simulations by incrementing in 16 steps the testing and tracking efficacy from 0 to 100%. For the purpose of the simulation, the testing and tracking strategy was assumed to be introduced 20 weeks after the first outbreak.

#### 2.2.2 Model parameters and latent causes

The model we used is formally identical to the one described in (Friston et al., 2020 c,d) and the full list of model parameters can be found in Table SM-1. Here, we describe in greater detail the hidden states that are particularly relevant for the present purposes (‘hidden’ means that these quantities were not directly observed but were *inferred* by the model):

- The *probability of leaving home* (Location factor) given the condition of being asymptomatic, has a prior baseline rate (see ‘probability of going out’ parameter in Table SM-1) multiplied by a decreasing function of the proportion of infected people. The social distancing, modeled as “the propensity to leave home and expose oneself to interpersonal contacts” [Friston et al 2020a]”, is an exponential threshold parameter (see social distancing threshold Table SM-1).
- The *proportion of infected people* (Infection factor) represents the proportion of people that are infected at a certain time. The probability of being infected depends upon the number of social contacts, and the proportion of time spent at home. These dependencies are parametrized by the “effective number of contacts being home and being out” and the “probability of getting contagion for each contact” (Table SM-1)
- The *proportion of immune people* (Infection factor) represents the amount of people that are probabilistically immune at a certain time.
- The *proportion of resistant people* (Infection factor) represents individuals that are resistant to virus exposure. They represent a portion of the effective population that are not susceptible to infection because they are relatively protected from the infection by immunity such as cross-reactivity [Grifoni et al 2020, Ng et al 2020] or protective hosts factors [Bunyavanich et al., 2020; Zheng et al., 2020]. Notably during the epidemic, people can transit from a state of exposure to a state of resistance through a mild illness that does not entail seroconversion (recovery is mediated by T-cell response [Chau et al., 2020]. The resistant state thus corresponds to the immune state for subjects who never became contagious.
- The *proportion of people showing symptoms* (Symptoms factor) represents the proportion of subjects developing symptoms after being exposed.

The results were obtained by using MATLAB code available as part of the free and open source academic software SPM (https://www.fil.ion.ucl.ac.uk/spm/), released under the terms of the GNU General Public License version 2 or later.

#### 2.2.3 Correlational analyses

In order to evaluate the capacity of the model of inferring latent variables, we assessed how well the probability of leaving home tracked the time series of cellphone movements in the latest period available (from February 1^th^ to March 27^th^), via a Pearson’s correlation analysis.

## 3 Results

### 3.1 Model validation and predictive validity

The time series of estimated daily number of positive cases and daily number of deaths, as compared with the actual data are reported in Fig 2–3. The predictive capability of the model was assessed by withholding data from the last 20 days during model inversion (see Methods). The time series considered included data from January 22^nd^ to June 30^th^. This choice reflects a peculiar social habit characterizing Italian population (a strong tendency to go on vacation in a period ranging from early July to late August). The model forecast was then compared to the actual data for these time points (Fig 2–3 insets; model forecast: black lines; actual data: red dots). If we used data up to July 20^th^ one can see that while the model predicted a slight increase for the number of daily positive cases—which is arguably due to starting of decaying the immunity— inaccurate predictions are most likely due to isolated local outbreaks typically occurring during vacation periods, people returning from foreign countries after vacations or migrants fluxes (for a full description of the effects of vacations see Supplementary Material). In all cases the data trends related to dataset 1 (see Methods) fall within the Bayesian confidence interval both for the positive cases and for the daily deaths.

**Fig. 3.**
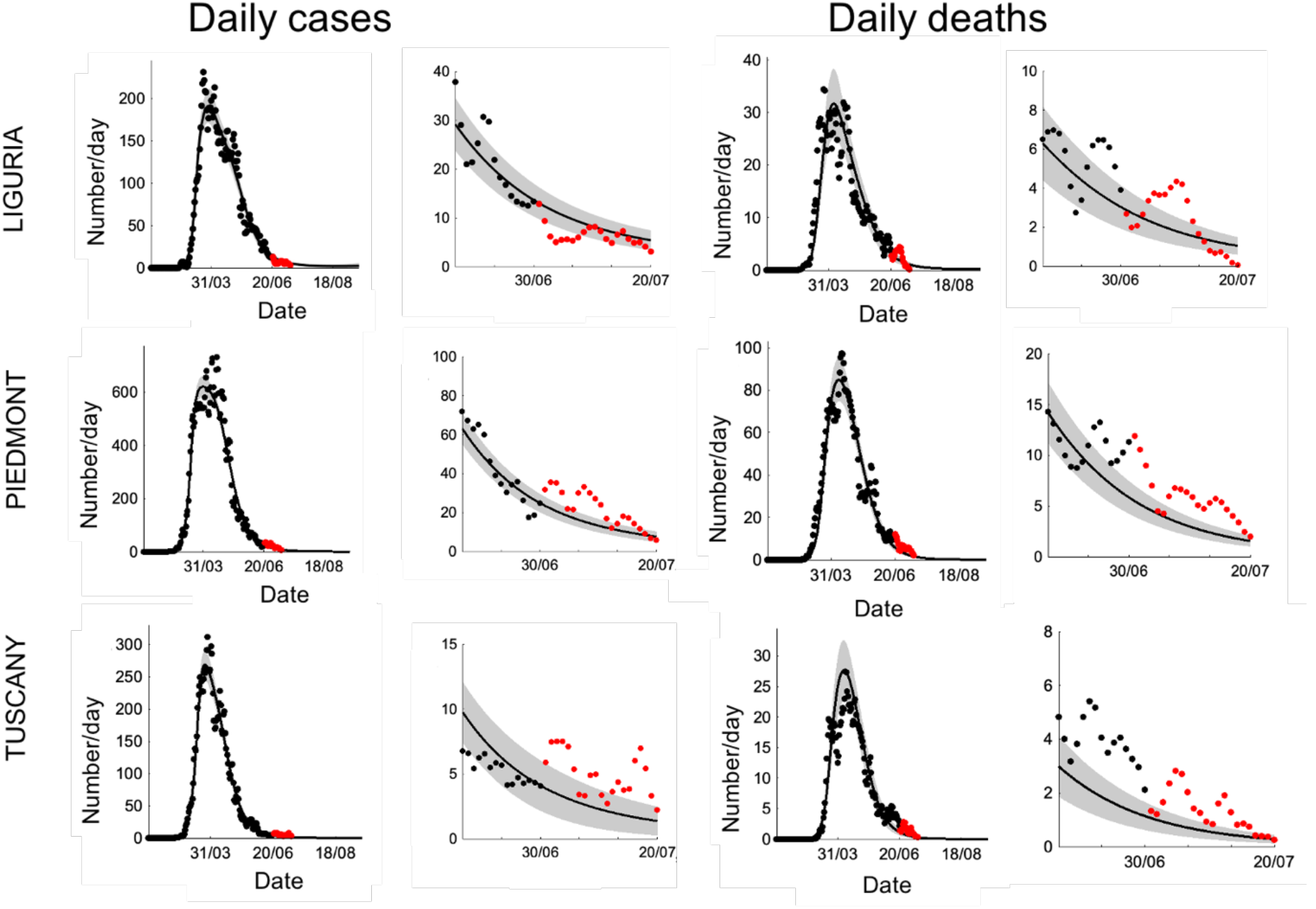
Predictive validity of the model. (Liguria, Piedmont and Tuscany). This figure uses the same format as figure 2.

#### Estimation of the duration of immunity and second-wave forecast

The Bayesian model comparison procedure, within a set of models with varying periods of immunity (PIL) from 1 to 32 months (see Methods), yielded a PIL best estimate of 7 months (Fig. SM-4a). This estimate was used to generate a long-term prediction for recurrences of COVID-19 epidemic in the different Italian regions. As shown in Fig. 4, a second wave is expected for all regions under consideration, although with different probability and intensity. The peaks of the second waves range from September-October 2020 (Veneto, Emilia-Romagna and Lombardy) to December 2020 (Piedmont). It should be noted that a 7-month PIL best estimate was also obtained with dataset 1 (data not shown), suggesting that this value is not significantly influenced by the recently observed outbreaks. Furthermore, both simulations predict the occurrence of second waves for all the six regions, variably peaking from late summer to early winter.

**Fig. 4.**
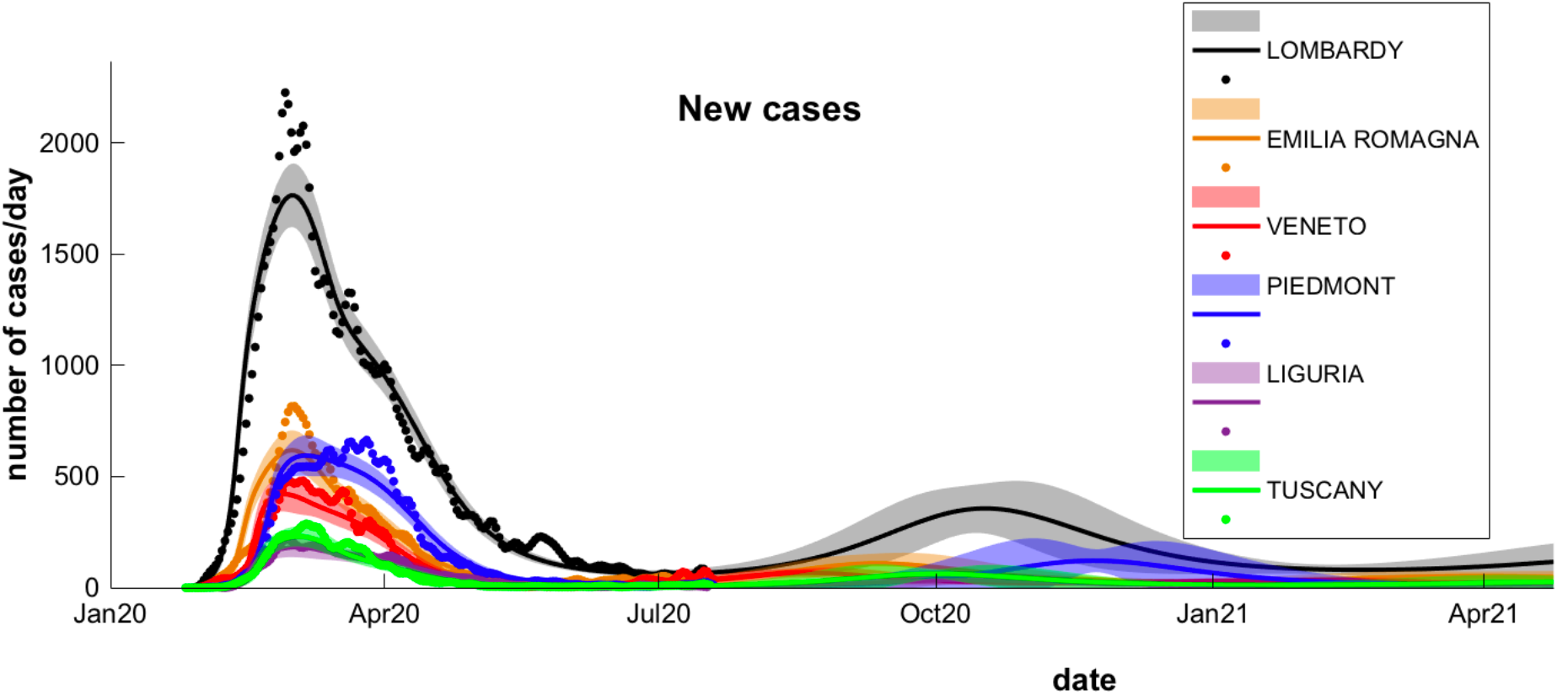
Long-term prediction following Bayesian model comparison. Comparison of data fitting and prediction for the six regions under consideration, obtained following a Bayesian model comparison of 32 models for each region. Dots (different colors for each region) represent data on daily positive cases reported by the “National Civil Protection Agency”. The lines reproduce posterior expectations (different color for each region) with the corresponding 90% Bayesian confidence bands (shaded). Note the different timings of the peak of the second wave and its relative intensity in the different regions.

#### Latent causes driving the pandemic

The salient factors determining the differences between regions were considered in some detail. The model was inverted using the daily number of tests available in the official repository of the “Italian Civil Protection”, together with daily number of positive cases and deaths. Additionally, on the basis of the Bayesian model comparison results, the prior estimate of the PIL was fixed to 7 months. The posterior estimates of this simulations are shown in Table 1.

Table 1

**Table1**
| FACTORS | PARAMETERS | LOMBARDY | VENETO | EMILIA - ROMAGNA | LIGURIA | PIEDMONT | TUSCANY |
| --- | --- | --- | --- | --- | --- | --- | --- |
| Location parameters | Prob going out from home (%) | 32.81 | 35.35 | 33.46 | 33.06 | 33.37 | 33.07 |
|  | Social distancing threshold | 0.0244 | 0.0297 | 0.0340 | 0.0302 | 0.0319 | 0.0265 |
|  | Bed availability threshold (per capita) | 0.0001671 | 0.0001667 | 0.000165 | 0.000167 | 0.000168 | 0.000168 |
| Infection parameters | Effective number of contacts being at home | 5.92 | 3.18 | 5.55 | 6.03 | 6.13 | 5.99 |
|  | Effective number of contacts being out | 51.58 | 99.78 | 55.39 | 42.05 | 45.44 | 54.34 |
|  | Probability of getting contagion for each contact (%) | 0.52 | 0.56 | 0.53 | 0.44 | 0.48 | 0.57 |
|  | Infected pre contagious period (days) | 3.94 | 3.35 | 3.80 | 3.71 | 3.66 | 3.21 |
|  | Infected contagious period (days) | 3.43 | 3.65 | 4.16 | 3.82 | 3.89 | 3.48 |
| Clinical parameters | Time till symptoms (days) | 16.71 | 19.61 | 18.35 | 17.89 | 18.81 | 21.41 |
|  | Prob severe symptoms from symptomatic conditions (%) | 2.87 | 2.56 | 2.79 | 2.76 | 2.66 | 2.26 |
|  | Symptomatic period (days) | 8.68 | 9.57 | 8.95 | 8.85 | 9.29 | 10.38 |
|  | CCU period (days) | 14.82 | 15.68 | 14.62 | 12.61 | 16.85 | 13.72 |
|  | Prob of death from CCU with severe symptoms (%) | 50.91 | 44.06 | 48.32 | 49.93 | 50.25 | 39.09 |
|  | Prob of survival from home with severe symptoms (%) | 12.61 | 12.56 | 12.59 | 12.65 | 12.71 | 12.56 |
| Testing parameters | Test, track and trace | 0.0001 | 0.0001 | 0.0001 | 0.0001 | 0.0001 | 0.0001 |
|  | Test delays (days) | 1.15 | 1.10 | 1.13 | 1.44 | 1.32 | 1.24 |
|  | Test selectivity (for infection) | 11.38 | 3.36 | 9.05 | 8.72 | 10.56 | 4.03 |
|  | Baseline testing | 0.029 | 0.074 | 0.064 | 0.057 | 0.023 | 0.034 |
| Immunity | Proportion of resistant cases (%) | 41.86 | 73.24 | 64.76 | 52.46 | 57.56 | 65.35 |
|  | Proportion of people with innate immunity (%) | 57.21 | 44.80 | 46.14 | 48.71 | 48.85 | 45.37 |

The model was able to fit data accurately (see Fig. SM-1) and to generate predictions of the daily deaths and positive cases over the long-term. Furthermore, the model was able to estimate many hidden parameters that underwrite epidemic dispersion. In Fig. 5 the probability of leaving home during the pandemic is shown for all regions under consideration. Interestingly, the probability of leaving home reflects changes in behavior, due to the lockdown strategies progressively imposed by the Italian government, which increased lockdown severity from February 23^rd^, when the initial so-called ‘soft’ lockdown was applied to the country, to March 8^th^, when a tight lockdown was first imposed in the most affected areas of northern Italy and, one day later, throughout the country.

Indeed, the model-predicted dynamics closely reflect the official lockdown dates (cfr Fig. 5). It should be noted that, beside a general common trend, Veneto showed a different behavior since the probability reached a shallower minimum compared to the other regions—and returned to the initial value within a few days. Therefore, Veneto’s parameters speak to less strict adherence to lockdown. A further model validation came from the correlation of cellphone movements (data limited to March 27^th^), with the probability of leaving home inferred by the model. The reduction of mobility inferred by the model was lower for Veneto compared to Lombardy and Emilia-Romagna (Fig. 6), matching the cellphone movements (see also Vinceti et al., 2020). The correlation of cellphone movements with the probability of leaving home — over the whole time series — ranged from a minimum of 0.85 for Emilia-Romagna and Veneto to a maximum of 0.97 for Piedmont (Pearson’s correlation coefficient;Fig. 6, Table 2).

**Fig. 5.**
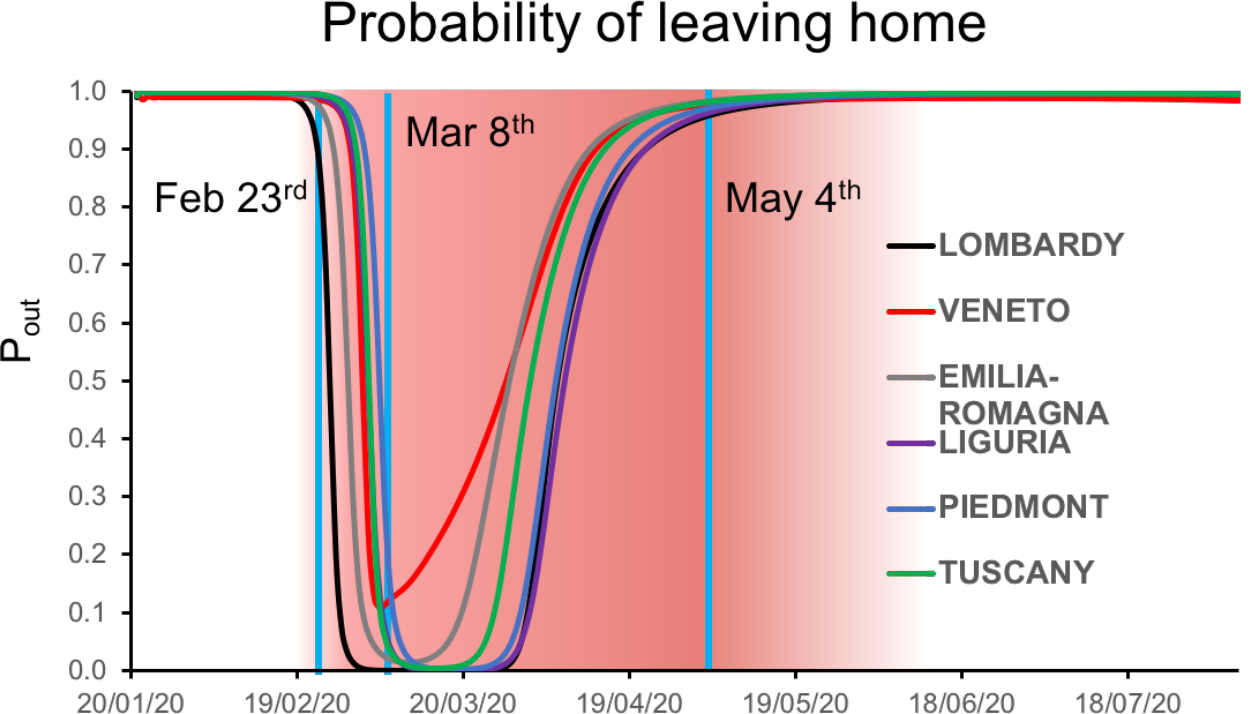
Dynamics of the probability of leaving home. Plot shows the kinetics of the probability of leaving home inferred by model inversion (colored lines) in the time window ranging from January 22^nd^ to August 22^nd^. During this period, the Italian government imposed a lockdown with different degrees of severity from February 23^rd^ (soft lockdown: country-wide closing of schools, universities, and all non-essential industrial and commercial activities; limiting the activities of public offices) to March 8^th.^ (tight lockdown: prohibition of any kind of mobility, apart from specific health or professional needs). May 4^th^ is the date where the tight lockdown was relaxed and people were allowed to freely move within their region. Vertical blue lines indicate the date of lockdown, while the shaded red region represents the lockdown time window. Note the peculiar behavior of Veneto (red line), showing a less intense reduction of the probability of leaving home, indicating a reduced adherence to lockdown policies.

**Fig. 6.**
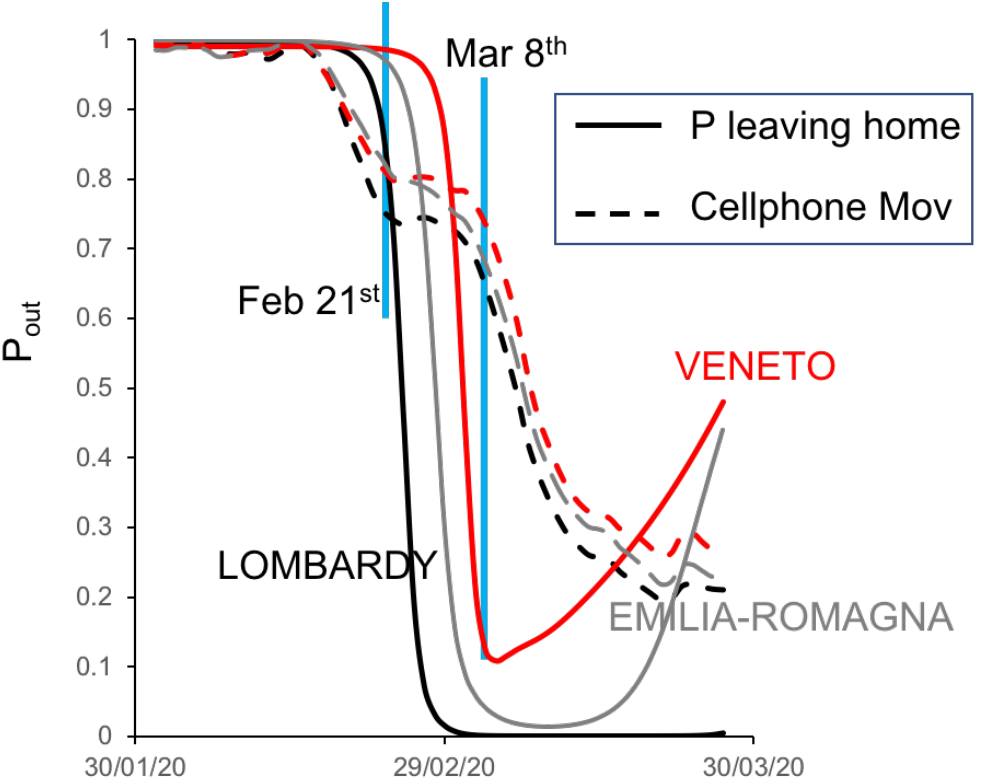
Dynamics of the probability of leaving home, compared with cellphone movements. Plot showing the trajectory of the probability of leaving home inferred by the model (solid lines) and the monitored cellphone movements (dashed lines) for the three most hit regions (Lombardy, Veneto and Emilia-Romagna) in the period from February 21^st^ to March 27^th^

**Table 2.** Pearson’s coefficient resulting from the correlation between the time series of cellphone movements and the probability of leaving home.

| *Pearson's coeff* |  |
| --- | --- |
| LOMBARDY | 0.87 |
| VENETO | 0.85 |
| EMILIA-ROMAGNA | 0.85 |
| LIGURIA | 0.94 |
| PIEDMONT | 0.97 |
| TUSCANY | 0.94 |

The latent causes generating the dynamics of the epidemic are related to one of the states in the model (LIST, see Methods). Focusing on infection factors, it is possible to infer the dynamics of the pandemic in terms of susceptibility, state of infection, immunity or forms of resistance to the virus. Fig. 7 shows the dynamics of three of the infection factors inferred by the model. As is evidenced by the plot (Fig. 7A), the proportion of infected people shows a rapid rise during the first weeks of the epidemic peaking variably from region to region, with Lombardy showing the highest and Veneto the lowest prevalence. Similarly, the proportion of immune people rises quickly in the first weeks of the epidemic peaking with different values from region to region, with Veneto and Lombardy at the lower and upper extremity of the range, respectively (Fig. 7B).

**Fig. 7.**
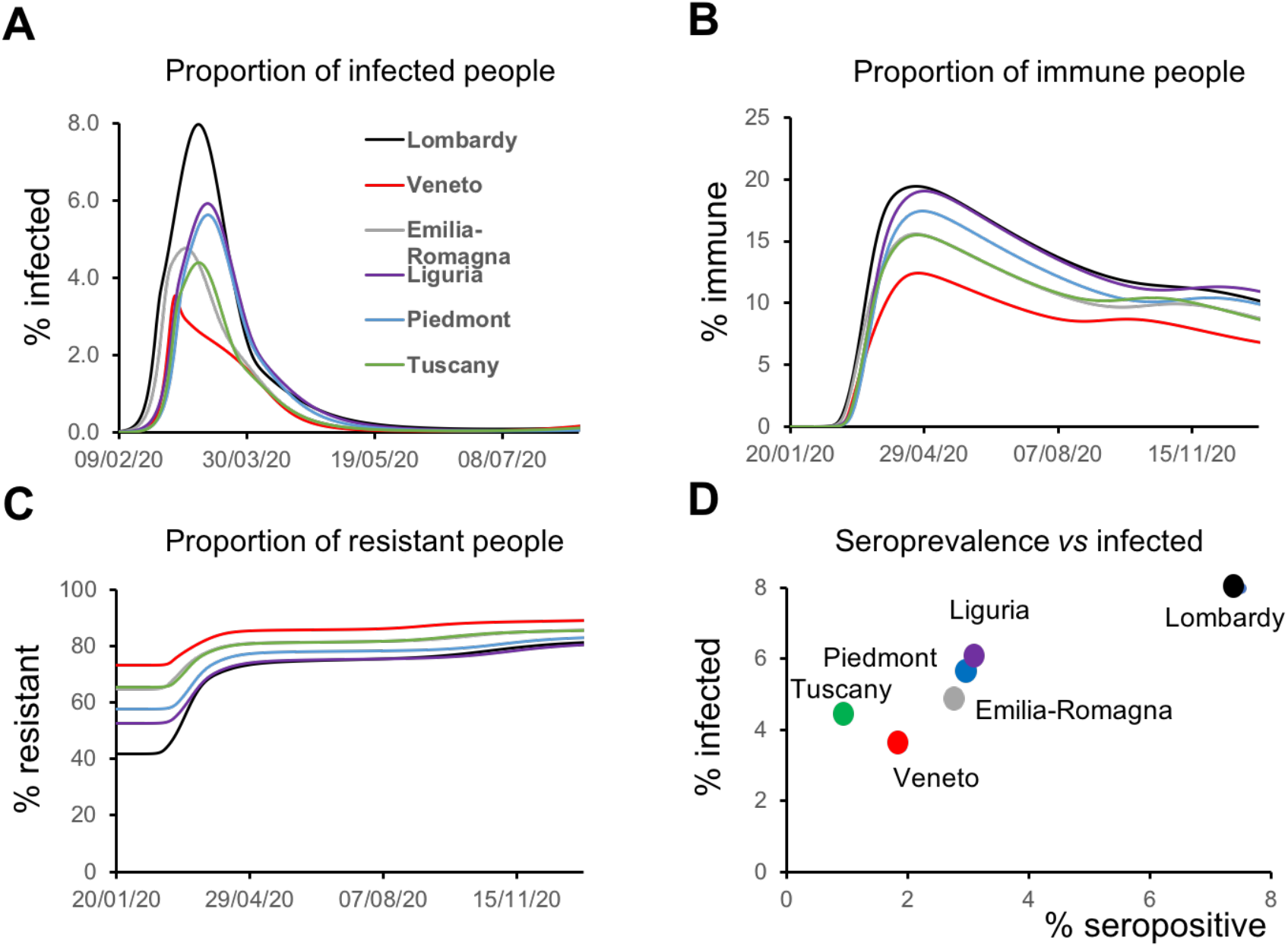
Latent causes. **A** Plot shows the trajectory of the proportion of infected people during the epidemic and in the following months for all the regions considered. Note that Veneto shows a steep rise at the beginning of the epidemic but quickly drops to the initial value, while other regions present slower kinetics. **B** Plot shows the kinetics of the proportion of immune people during the epidemic and in the subsequent months for all the regions considered. As in the case of infected people, the proportion of immune people is less in Veneto throughout the considered time window **C** Plot shows the trajectory of the proportion of resistant people during the epidemic and in the subsequent months for all the regions considered. Here, the proportion of resistant people is higher in Veneto at the beginning of the epidemic. **D** Data on serological test (% of population that have been infected) are plotted against the peak value of the proportion of infected people inferred by the model and shown in panel A.

As immunity is lost, the susceptible proportion gently rises back towards the initial value. However, the proportion of resistant (see Methods) subjects rises in response to the accumulation of people recovering from illness and therefore leaving the susceptible reservoir (Fig. 7C). At the end of May, the Italian government launched a screening campaign to randomly sample the population, in order to test whether they had been infected by coronavirus. As can be seen in Fig. 7D, the proportion of infected people inferred by the model matches the results of the serological tests (see Methods), in all the regions under consideration (R^2^ = 0.92 with linear regression). Furthermore, i) the ranking of the regions in terms of population infected is identical in both cases; ii) the order of magnitude of the proportion of seropositive and infected is comparable and, in some cases (e.g. Lombardy), the numerical values are identical. This is a remarkable aspect of predictive validity because these serological data where never used to inform the model parameters.

#### Second wave forecasts and future scenarios

Given the tight matching between the latent causes inferred by the model and empirical measures of lockdown, social mobility and seroprevalence, the model was used to generate long and mid-term predictions. The posterior parameter estimates (Table 1) predict the presence of a second wave of infection in all the considered regions. Notably, the time of the second waves varies from mid-September to late November.

(Fig 8–9, red lines). This forecasting is tightly coupled to the recurrence of infections observed in recent weeks (Fig 2–3 red dots), related to isolated outbreaks spotted throughout Italy. The early peaks are in fact predicted in Veneto (mid-September) and Tuscany (early October), regions that witnessed an unexpected rise in the number of positive cases (see Fig. SM-2 and SM-3). Interestingly, the time of the second wave for these regions is significantly delayed if data related to these increases are withheld from model inversion, indicating the capacity of the model to continuously improve its predictions as new data become available (especially data containing information about novel relevant events, such as the sharp changes in people movements in late July-August, in the case of Italy).

**Fig. 8.**
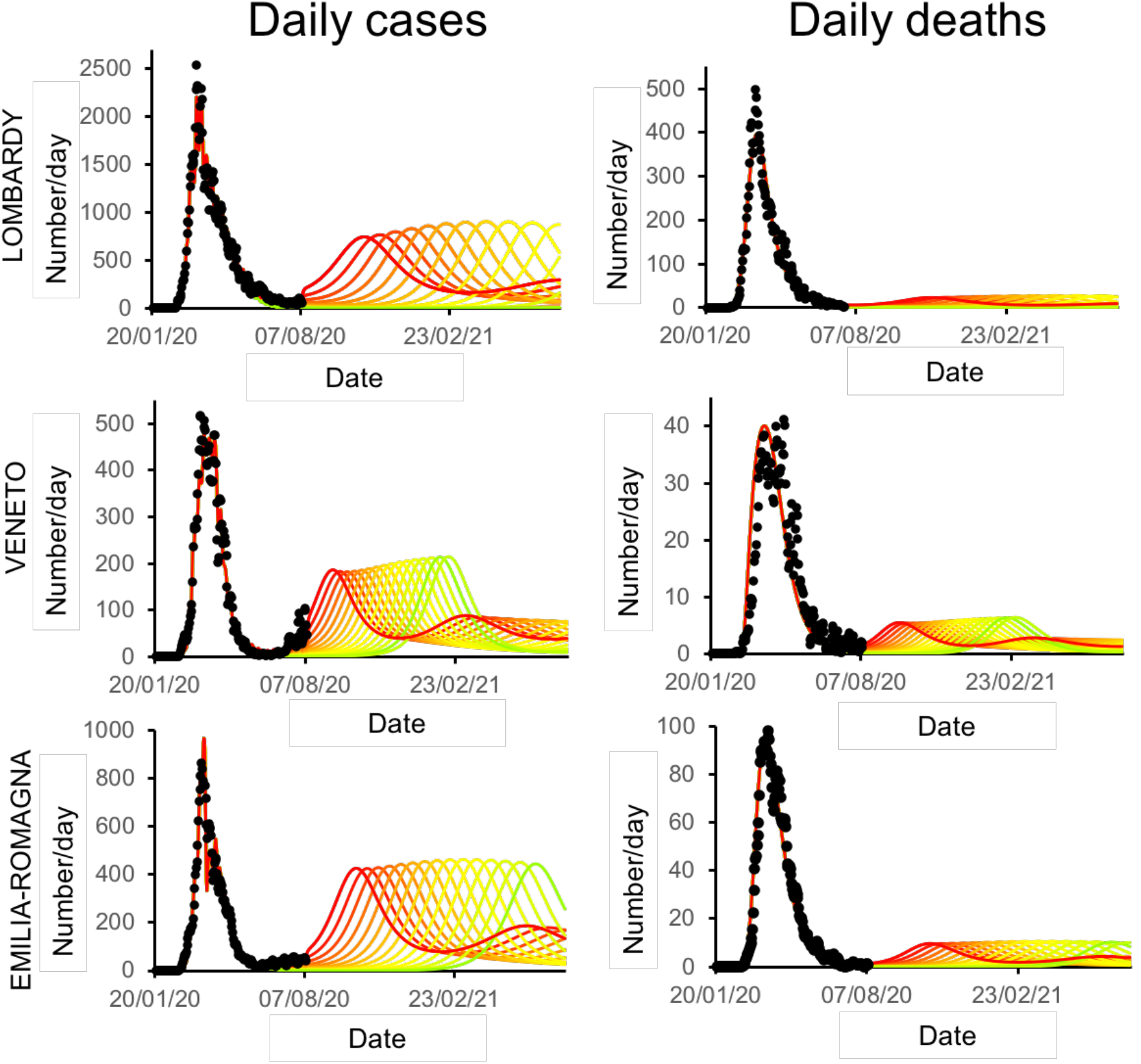
Second-wave forecasts (Lombardy, Veneto and Emilia-Romagna): Left. Plots show the effect of increasing the efficacy of testing and tracking strategies on the time and peak amplitude of the second wave for daily positive cases. These trajectories go from zero (red line) to 100% efficacy (green line). **Right** Similarly, the plots in the right column report predictions of daily deaths under increasing levels of testing and tracking efficacy.

**Fig. 9.**
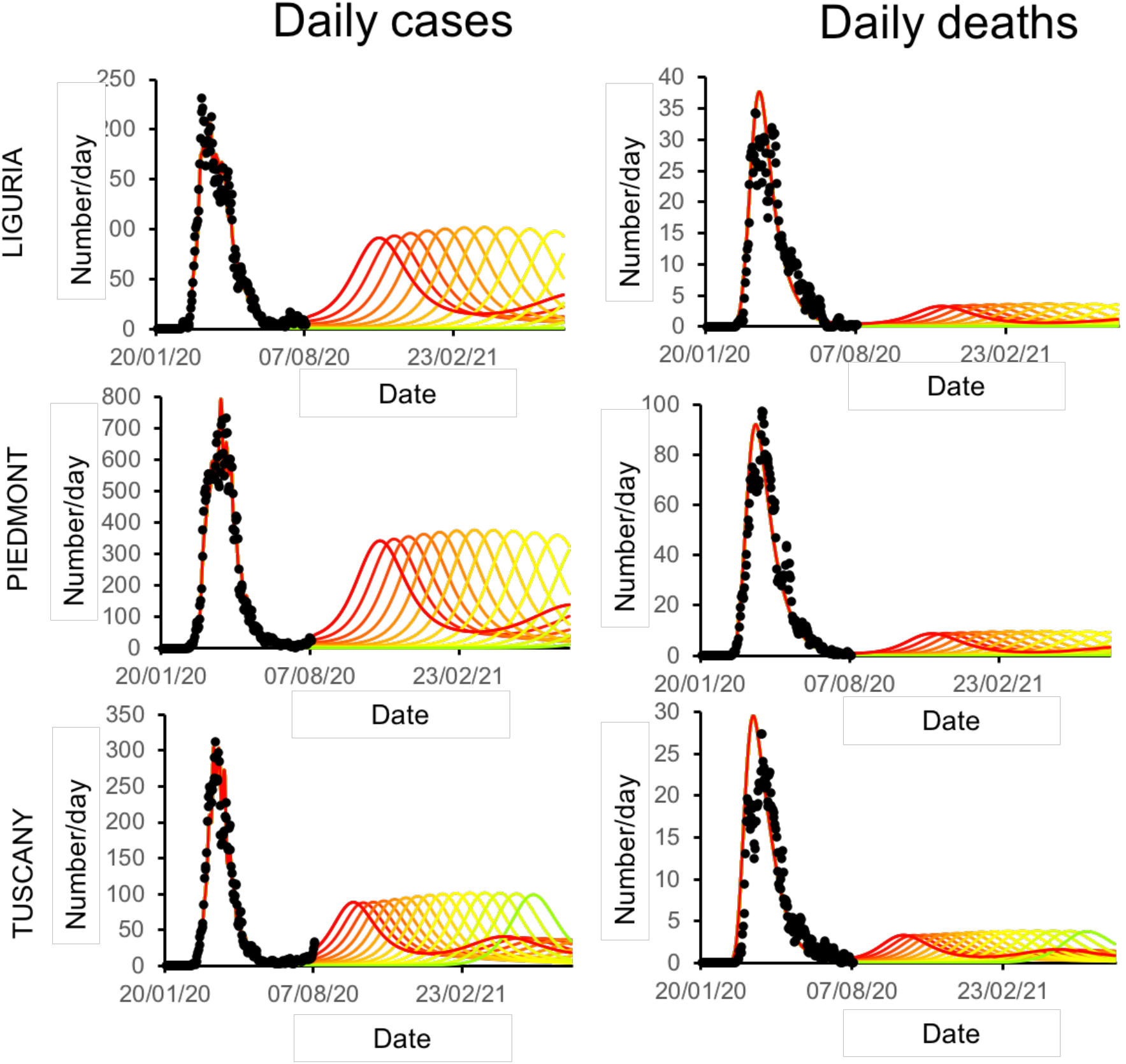
Second-wave forecasts (Liguria, Piedmont, Tuscany): Left. Plots show the effect of increasing the efficacy of testing and tracking strategies on the time and peak amplitude of the second wave for daily positive cases. The format of this figure follows that of the previousFigure 8

One common metric of viral transmission is the reproduction ratio (Rt), which is the estimate of the number of people infected by one person. This quantity can be inferred by the model. The effective reproduction rates for Italian regions reflect the lockdown policies that mitigated viral transmission through an efficient reduction of the Rt (Fig SM-5). Although the reproduction rate has remained below 1 for the period of the lockdown, during late June and July the model inferred an increase above 1 for all six regions, which has been confirmed by current data released by the Italian Minister of Health.

Finally, we analyzed how the efficacy of testing and tracing strategies could mitigate the spread of SARS-CoV-2 using the posterior estimates above. One of the most effective mitigation (and possibly suppression) strategies is to test for infected but asymptomatic subjects and trace their contacts to contain novel outbreaks [Alfano et al 2020, Kraemer et al 2020, Tobías 2020, Aleta et al 2020]. One can evaluate alternative contact tracing scenarios by simulating the future following an increase in the efficacy of testing and tracking.

In particular, the efficacy of testing and tracing strategy was increased — from the level initially inferred by the model — to the full efficacy (i.e. testing and tracing of *every* asymptomatic individual). As seen in Fig 8 and 9 (and summarized in Fig SM-6), the ensuing predictions for all regions show the occurrence of a second wave in the coming autumn with a significant reduction of prevalence (Fig 8,9 from red to green lines) and fatality (Fig 8–9 from red to green lines), as the testing and tracking efficiency is increased. Notably, the occurrence of a second wave can be postponed by a suitable enhancement of testing and tracing policies, even beyond the time window under consideration but such strategy can also markedly reduce the total number of deaths and positive cases (Figure SM-6). Moreover, the unexpected events observed in Veneto and Tuscany, giving rise to an anticipated second wave in the beginning of autumn, if properly treated could remain contained – thus avoiding a putative second wave.

The posterior estimates were analyzed to understand whether the observed differences among regions under consideration could be attributed to some of the factors/parameters included in the model. As seen in Table 1, the parameters related to testing factor show peculiar characteristics for Veneto, which exhibits a low selectivity for testing the infection (almost half of the other regions) and a very high baseline testing. Furthermore, Veneto also displays a significant proportion of resistant cases, which means that most of the population in that region may have some distinctive characteristics related to personal, environmental and most probably social behaviors. Indeed, Veneto implemented a different testing policy since the early begin of the outbreak, by testing both symptomatic and asymptomatic subjects, while in other regions only symptomatic cases were investigated [Romagnani et al 2020, Binkin et al 2020]. This different testing policy seemed to have reduced the virus spread in that region --- as reflected in the lower pandemic toll compared to other regions --- due to identification of both documented and undocumented infections, the latter ones being the main source of the documented cases, accounting up to 80% [Li et al 2020] of the infections.

## Discussion

Following the first few dramatic weeks of the COVID-19 pandemic, in which the countries and the healthcare system had to face an unprecedented emergency, attention is now turning to the hidden causes of the marked differences in COVID-19 spread from country to country and region to region, with the aim of trying to predict the future course of the pandemic and to favor the adoption of adequate public health measures to curb the outbreak. The DCM approach differs from other mathematical models because of its commitment to a generative model that contains all the latent factors responsible for the epidemic—and for generating empirical timeseries. Similar to brain dynamics, in which the activity of single neurons and neuronal populations can be characterized in detail with sparse and noninvasive data, the individuals and communities playing a role in a pandemic can be characterized, using aggregated data on morbidity and mortality at a population level. DCM, which was originally conceived to characterize complex multiscale systems, was able to infer several epidemiologically parameters subtending the progression of the pandemic.

### Hidden factors driving COVID-19 diffusion in Northern Italy

Among the parameters estimated by the model, the effective population, representing the number of people that are affected by the outbreak, was smaller than the total (census) population size and the effective proportion of the total population varied over the different regions (Table SM-3). For instance, Tuscany had only 20%, while Lombardy, one of the most severely affected European regions, had almost 90% of the population involved. This value, which provides an estimate of the overall impact of COVID-19 on the population, could indeed reflect different geospatial factors, the territory, lifestyle, behavior and social habits. For instance, Lombardy has more than twice the number of towns with > 20K inhabitants compared to Tuscany, which could favor aggregation and viral spread, especially in the initial phase of the pandemic.

Infection parameters were similar in the 6 regions under consideration [Lauer et al 2020, Ortiz-Prado et al 2020] supporting the fact that virology is substantially equal, despite potential differences in climate and other geographic factors. Likewise, location and clinical parameters were similar in the 6 regions indicating that the actuation of social and sanitary healthcare strategies did not differ remarkably. It is relevant that the model highlights differences in the testing parameters of Veneto. This evidence is in line with the fact that the prevention policies—realized by local administrators in that region—were effective in containing viral spread. An even more striking prediction pertains to effective immunity. The proportion of population with innate immunity falls around similar values (45%) for all regions, except Lombardy, showing that more than half of the population (57%) in that region were refractory to the infection. This may reflect a higher initial level of immunity determined by the earlier circulation of the coronavirus, whose presence was detected before the first reported case [Zehender et al 2020].

Another interesting estimate is the proportion of resistant subjects, which was lower in Lombardy (42%) compared to Veneto (73%) and Emilia Romagna (65%). Taken together, parameter estimates regarding the susceptible population and innate immunity substantiate the highest impact of COVID-19 epidemic in Lombardy compared to Veneto and Emilia-Romagna that were simultaneously hit by the outbreaks. These, in turn, may reflect a combination of causes including geographical segregation of the population, lifestyle, social habits — and environmental factors such as air pollution or climate conditions — that may favor the virus persistence and thus individual exposure [Vinceti et al 2020, Filippini et al 2020].

### Model validation

The model was validated by comparing the available epidemiological data with forecasts generated by simulations. The changes in location states during the acute phase of the pandemic closely matched the lockdown enforced by the government (DCPM 1,2,3,4). Notably, the model-inferred changes in mobility as a consequence of the lockdown were validated by comparison with data on actual mobility, as tracked by cellphone movements. More importantly, the model-inferred probability of being infected during the epidemic, reflecting the proportion of infected people, was strongly correlated with the data from serological tests promoted by the Italian government.

The model was also validated through a procedure typical in machine learning (see predictive ability in the Methods section). There is good agreement between model and data when considering the input time series limited to the period from January to June 30^th^, while some points (positive cases) fall outside the confidence intervals when considering the input time series extended to July 23^rd^. It is important to note that, after June 30, sudden infection foci were related to episodic and isolated outbreaks in confined areas. This may be explained as follows: i) Traditionally, the Italian population moves around during late July and August for vacation. ii) Italy is traditionally one of the preferred destinations for international tourism and, although this year it is markedly reduced, the country remains exposed to imported cases. iii) Most of the cases observed in the first days of August are related to Italian tourists coming back from foreign countries, in which the infections are increasing. iv) In summer, Italy is particularly exposed to migrants, and this year is no exception. Migrants, when intercepted, are immediately screened and quarantined but some probably escape the entry filter and propagate the infection.

Differently from what was reported in (Friston et al 2020d), where the immunity period loss for worldwide countries was estimated in three months, we obtained a longer period (seven months), which could be due to the intrinsic characteristic of the pandemic evolution. Indeed, the curve of new cases in the Italian regions peaked earlier than curves for other countries such as USA or Brazil, where the pandemic outbreak after more than 5 months is still in tumultuous evolution. Furthermore, the original reports of three months were based upon earlier analyses of shorter time series. This produced larger confidence intervals. It is likely that as more data becomes available, the effective period of immunity may increase even further.

### Second wave prediction and alternative scenarios

As the reliability of the model has been demonstrated by several validation procedures, we provided predictions for the COVID-19 pandemic in the upcoming months in the geographical areas under study. Particular attention should be paid to the differences in forecasting resulting from the simulation with data up to the end of June, compared to the one with the data including the first days of August (Fig. 2–3 vs Fig. SM-2 and SM-3). Among the investigated regions, Lombardy shows small differences in the predicted number of cases and deaths, while Veneto, Piedmont and Tuscany show a sharp increase in the first week of August that was not predicted when the model inversion was performed on the data up to the end of June. Indeed, these new cases are mainly related to an increase of population fluxes among regions, returning from vacations or alternatively, because of immigrating fluxes. These unforeseen events cannot be predicted by the model, nevertheless since the sudden rise of the curve can portend a second wave, the model adjust its prediction accordingly. Notably, the rising forecast for the last week of July and the first week of August — which may eventually lead to a second wave — could reflect a synergetic effect due to the temporal coincidence of immunity decay and the sharp increase in the vacation and migration fluxes (the end of July is 7 months past the end of January). Furthermore, the evaluation of the impact of different strategies of healthcare policy (Fig. 8) — mainly by increasing the efficacy of testing and tracking (see Methods) — shows they can nuance the second wave and reduce its impact in terms of lives saved and pressure on the healthcare system. In other words, while a second wave is extremely plausible, it can be postponed – if properly contained – and remain of limited intensity. Nonetheless, in all cases, the severity of the second wave predicted by the model is markedly lower than the first wave in terms of deaths, proportion of infected people and proportion of people showing symptoms (see Table SM-2).

### Model strength and limitations

The proposed study, despite the advantages given by DCM, suffers from some limitations, which have already been mentioned in [Friston et al. 2020d]. i) The DCM does not consider interactions with seasonal flu or other annual fluctuations [Kissler et al 2020]. ii) As with other modeling approaches, the outcomes of the Bayesian model comparison and posterior inferences are strictly model-dependent. iii) Updating the model with available data could change the posterior predictions. iv) The model does not include geospatial aspects, but rather each outbreak is treated as a point process [Chinazzi et al 2020].

Nevertheless, i) annual fluctuations like seasonal flu can alter the overall statistics but are normally distributed throughout the country without preferential geographical localizations, therefore the eventual error is systematic and homogeneously distributed; ii) the outcomes of every model are limited to the approach that is actually employed; iii-iv) the continuous updating of the model and therefore of the posterior predictions is inherently one of the main advantage of this kind of approaches. The flexibility of the model takes into account unexpected and sudden events that can promptly change the pandemic dynamics allowing therefore to continuously update the prediction generating different future scenarios; iv) once more, treating each outbreak as a point process provides the model with the capability to interpret single and spotted events as events potentially dangerous, rather than needing to collect all the data to yield a uniform prediction.

### Conclusion and implications

This work presents several innovative aspects: i) at a scientific level, the approach proposed by Friston et al—of applying the method originally developed to investigate brain functions to the COVID-19 pandemic—is undoubtedly innovative and its performance was remarkable in terms of fitting and predictive power. Additionally, the retrospective analysis of the hidden factors underlying the pandemic could furnish an innovative epidemiological tool. ii) The DCM approach enables the evaluation of the role of factors that can be manipulated by institutional and healthcare policies. iii) The preventive examination of the possible scenarios of second waves, according to different choices of healthcare policy, allows a better planning by optimizing the benefit-cost ratio in view of the specific means and resources available to each region. In other words, analyzing the different scenarios predicted by the model should allow regional authorities to balance complimentary strategies, like lockdown or testing-and-tracking, allowing them to minimize the impact on the sanitary and economic system.

The good performance of these Italian regions (and of Italy as whole) compared to other neighboring countries may derive from the severe lockdown and the maintenance of social distancing and prevention strategies. Thus, while waiting for effective vaccines, the second wave could be reduced and diluted by maintaining and enhancing the prevention strategies currently in use.

## Data Availability

Data will be made available upon request

## Footnotes

1 http://www.salute.gov.it/portale/news/p3_2_1_1_1.jsp?lingua=italiano&menu=notizie&p=dalministero&id=4998;_0_file.pdf

## References

1) Vinceti M, Filippini T, Rothman KJ, Ferrari F, Goffi A, Maffeis G, Orsini N. (2020) “Lockdown timing and efficacy in controlling COVID-19 using mobile phone tracking” eClinicalMedicine. Open Access Published: July 13, 2020 DOI: https://doi.org/10.1016/j.eclinm.2020.100457

2) Bonaccorsi G, Pierri F, Cinelli M, Flori A, Galeazzi A, Porcelli F, Schmidt AL, Valensise CM, Scala A, Quattrociocchi W, Pammolli F. (2020). “Economic and social consequences of human mobility restrictions under COVID-19.” Proc Natl Acad Sci U S A. 2020 Jul 7;117(27):15530–15535. doi: 10.1073/pnas.2007658117. Epub 2020 Jun 18.

3) Di Domenico L, Pullano G, Sabbatini CE, Boelle PY, Colizza V. (2020) “Impact of lockdown on COVID-19 epidemic in Ile-de-France and possible exit strategies.” BMC Med. 18(1):240. doi: 10.1186/s12916-020-01698-4.

4) Kucharski AJ, Klepac P, Conlan AJK, Kissler SM, Tang ML, Fry H, Gog JR, Edmunds WJ; CMMID COVID-19 working group. (2020) “Effectiveness of isolation, testing, contact tracing, and physical distancing on reducing transmission of SARS-CoV-2 in different settings: a mathematical modelling study.” Lancet Infect Dis. 2020 Jun 15:S1473-3099(20)30457–6. doi: 10.1016/S1473-3099(20)30457-6. Online ahead of print.

5) Metcalf CJE, Morris DH, Park SW. (2020) “Mathematical models to guide pandemic response.” Science. 2020 Jul 24;369(6502):368–369. doi: 10.1126/science.abd1668.

6) Van Kleef E, Robotham JV, Jit M, Deeny SR, Edmunds WJ. (2013) “Modelling the transmission of healthcare associated infections: a systematic review.” BMC Infect Dis. 2013 Jun 28;13:294. doi: 10.1186/1471-2334-13-294.

7) Chowell G, Sattenspiel L, Bansal S, Viboud C. (2016). “Modelling the transmission of healthcare associated infections: a systematic review.” Phys Life Rev. 2016 Sep;18:66–97. doi: 10.1016/j.plrev.2016.07.005. Epub 2016 Jul 11.

8) Paiva HM, Afonso RJM, de Oliveira IL, Garcia GF. (2020) “A data-driven model to describe and forecast the dynamics of COVID-19 transmission.” PLoS One. 2020 Jul 31;15(7):e0236386. doi: 10.1371/journal.pone.0236386. eCollection 2020.

9) Hauser A, Counotte MJ, Margossian CC, Konstantinoudis G, Low N, Althaus CL, Riou J. (2020) “Estimation of SARS-CoV-2 mortality during the early stages of an epidemic: A modeling study in Hubei, China, and six regions in Europe.” PLoS Med. 2020 Jul 28;17(7):e1003189. doi: 10.1371/journal.pmed.1003189. eCollection 2020 Jul.

10) Karnakov P, Arampatzis G, Kičić I, Wermelinger F, Wälchli D, Papadimitriou C, Koumoutsakos P. (2020) “Data-driven inference of the reproduction number for COVID-19 before and after interventions for 51 European countries.” Swiss Med Wkly. 2020 Jul 10;150:w20313. doi: 10.4414/smw.2020.20313. eCollection 2020 Jul 13.

11) Maugeri A, Barchitta M, Battiato S, Agodi A. (2020) “Modeling the Novel Coronavirus (SARSCoV-2) Outbreak in Sicily, Italy.” Int J Environ Res Public Health. 2020 Jul 9;17(14): E4964. doi: 10.3390/ijerph17144964.

12) Koopman J. (2004) “Modeling infection transmission. Annu Rev Public Health. 2004;25:303–26. doi: 10.1146/annurev.publhealth.25.102802.124353.

13) (a) Friston KJ, Parr T, Zeidman P, Razi A, Flandin G, Daunizeau J, Hulme OJ, Billig AJ, Litvak V, Moran RJ, Price CJ, Lambert C (2020) “Dynamic causal modelling of COVID-19” arXiv:2004.04463 [q-bio.P]

14) (b) Friston KJ, Parr T, Zeidman P, Razi A, Flandin G, Daunizeau J, Hulme OJ, Billig AJ, Litvak V, Moran RJ, Price CJ, Lambert C (2020) “Second waves, social distancing, and the spread of COVID-19 across America.” arXiv:2004.13017 [q-bio.PE]

15) (c) Friston KJ, Parr T, Zeidman P, Razi A, Flandin G, Daunizeau J, Hulme OJ, Billig AJ, Litvak V, Moran RJ, Price CJ, Lambert C (2020) “Testing and tracking in the UK: a dynamic causal modelling study.” arXiv:2005.07994 [q-bio.QM]

16) (d) Friston KJ, Parr T, Zeidman P, Razi A, Flandin G, Daunizeau J, Hulme OJ, Billig AJ, Litvak V, Moran RJ, Price CJ, Lambert C (2020) “Effective immunity and second waves: a dynamic causal modelling study.” arXiv:2006.09429 [q-bio.PE]

17) Friston, K.J., Harrison, L., Penny, W., 2003. Dynamic causal modelling. NeuroImage 19, 1273–1302.

18) Amato L, Fusco D, Acampora A, Bontempi K, Rosa AC, Colais P, Cruciani F, D’Ovidio M, Mataloni F, Minozzi S, Mitrova Z, Pinnarelli L, Saulle R, Soldati S, Sorge C, Vecchi S, Ventura M, Davoli M. (2020). “Volume and health outcomes: evidence from systematic reviews and from evaluation of hospital data.” Epidemiol Prev. 2017 Sep-Dec;41(5–6 (Suppl 2)):1–128. doi: 10.19191/EP17.5-6S2.P001.100.

19) Grifoni A, Weiskopf D, Ramirez SI, Mateus J, Dan JM, Moderbacher CR, Rawlings SA, Sutherland A, Premkumar L, Jadi RS, Marrama D, de Silva AM, Frazier A, Carlin AF, Greenbaum JA, Peters B, Krammer F, Smith DM, Crotty S, Sette A. (2020) “Targets of T Cell Responses to SARSCoV-2 Coronavirus in Humans with COVID-19 Disease and Unexposed Individuals.” Cell. 2020 Jun 25;181(7):1489–1501.e15. doi: 10.1016/j.cell.2020.05.015. Epub 2020 May 20.

20) Ng K, Faulkner N, Cornish G, Rosa A, Earl C, Wrobel A, Benton D, Roustan C, Bolland W, Thompson R, Agua-Doce A, Hobson P, Heaney J, Rickman H, Paraskevopoulou S, Houlihan CF, Thomson K, Sanchez E, Shin GY, Spyer MJ, Walker PA, Kjaer S, Riddell A, Beale R, Swanto C, Gandhi S, Stockinger B, Gamblin S, McCoy LE, Cherepanov P, Nastouli E, Kassiotis G, (2020). “Preexisting and de novo humoral immunity to SARS-CoV-2 in humans”. bioRxiv, 2020.2005.2014.095414.

21) Bunyavanich S, Do A, Vicencio A, (2020). “Nasal Gene Expression of Angiotensin-Converting Enzyme 2 in Children and Adults”. JAMA.

22) Zheng M, Gao Y, Wang G, Song G, Liu S, Sun D, Xu Y, Tian Z. (2020). “Functional exhaustion of antiviral lymphocytes in COVID-19 patients”. Cellular & Molecular Immunology 17, 533–535.ù

23) Chau NVV, Thanh Lam V, Thanh Dung N, Yen LM, Minh NNQ, Hung LM, Ngoc NM, Dung NT, Man DNH, Nguyet LA, Nhat LTH, Nhu LNT, Ny NTH, Hong NTT, Kestelyn E, Dung NTP, Xuan TC, Hien TT, Thanh Phong N, Tu TNH, Geskus RB, Thanh TT, Thanh Truong N, Binh NT, Thuong TC, Thwaites G, Tan LV, group, O.C.-r. (2020). The natural history and transmission potential of asymptomatic SARS-CoV-2 infection. Clinical Infectious Diseases.

24) Alfano V, Ercolano S. (2020) “The Efficacy of Lockdown Against COVID-19: A Cross-Country Panel Analysis.” Appl Health Econ Health Policy. 2020 Aug;18(4):509–517. doi: 10.1007/s40258-020-00596-3

25) Kraemer MUG, Yang CH, Gutierrez B, Wu CH, Klein B, Pigott DM; Open COVID-19 Data Working Group, du Plessis L, Faria NR, Li R, Hanage WP, Brownstein JS, Layan M, Vespignani A, Tian H, Dye C, Pybus OG, Scarpino SV. (2020) “The effect of human mobility and control measures on the COVID-19 epidemic in China.” Science. 2020 May 1;368(6490):493–497. doi: 10.1126/science.abb4218. Epub 2020 Mar 25.

26) Tobías A. (2020) “Evaluation of the lockdowns for the SARS-CoV-2 epidemic in Italy and Spain after one month follow up.” Sci Total Environ. 2020 Jul 10; 725:138539. doi: 10.1016/j.scitotenv.2020.138539. Epub 2020 Apr 6.

27) Aleta A, Martín-Corral D, Pastore Y Piontti A, Ajelli M, Litvinova M, Chinazzi M, Dean NE, Halloran ME, Longini IM Jr, Merler S, Pentland A, Vespignani A, Moro E, Moreno Y. (2020) “Modelling the impact of testing, contact tracing and household quarantine on second waves of COVID-19.” Nat Hum Behav. 2020 Aug 5. doi: 10.1038/s41562-020-0931-9. Online ahead of print.

28) Romagnani P, Gnone G, Guzzi F, Negrini S, Guastalla A, Annunziato F, Romagnani S, De Palma R. (2020) “The COVID-19 infection: lessons from the Italian experience.” J Public Health Policy. 2020 Sep;41(3):238–244. doi: 10.1057/s41271-020-00229-y.

29) Binkin N, Salmaso S, Michieletto F, Russo F (2020) “Protecting our health care workers while protecting our communities during the COVID-19 pandemic: a comparison of approaches and early outcomes in two Italian regions” MedrXiv doi: https://doi.org/10.1101/2020.04.10.20060707

30) Li R, Pei S, Chen B, Song Y, Zhang T, Yang W, Shaman J. (2020) “Substantial undocumented infection facilitates the rapid dissemination of novel coronavirus (SARS-CoV-2).” Science. 2020 May 1;368(6490):489–493. doi: 10.1126/science.abb3221. Epub 2020 Mar 16.

31) Lauer SA, Grantz KH, Bi Q, Jones FK, Zheng Q, Meredith HR, Azman AS, Reich NG, Lessler J. (2020) “The Incubation Period of Coronavirus Disease 2019 (COVID-19) From Publicly Reported Confirmed Cases: Estimation and Application.” Ann Intern Med. 2020 May 5;172(9):577–582. doi: 10.7326/M20-0504. Epub 2020 Mar 10.

32) Ortiz-Prado E, Simbaña-Rivera K, Gómez-Barreno L, Rubio-Neira M, Guaman LP, Kyriakidis NC, Muslin C, Jaramillo AMG, Barba-Ostria C, Cevallos-Robalino D, Sanches-SanMiguel H, Unigarro L, Zalakeviciute R, Gadian N, López-Cortés A. (2020) “Clinical, molecular, and epidemiological characterization of the SARS-CoV-2 virus and the Coronavirus Disease 2019 (COVID-19), a comprehensive literature review.” Diagn Microbiol Infect Dis. 2020 Sep;98(1):115094. doi: 10.1016/j.diagmicrobio.2020.115094. Epub 2020 May 30.

33) Zehender G, Lai A, Bergna A, Meroni L, Riva A, Balotta C, Tarkowski M, Gabrieli A, Bernacchia D, Rusconi S, Rizzardini G, Antinori S, Galli M. (2020) “Genomic characterization and phylogenetic analysis of SARS-COV-2 in Italy.” J Med Virol. 2020 Mar 29:10.1002/jmv.25794. doi: 10.1002/jmv.25794. Online ahead of print.

34) Filippini T, Rothman KJ, Goffi A, Ferrari F, Maffeis G, Orsini N, Vinceti M. (2020) “Satellite-detected tropospheric nitrogen dioxide and spread of SARS-CoV-2 infection in Northern Italy.” Sci Total Environ. 2020 Jun 16;739:140278. doi: 10.1016/j.scitotenv.2020.140278. Online ahead of print

35) Kissler SM, Tedijanto C, Goldstein E, Grad YH, Lipsitch M. (2020) “Projecting the transmission dynamics of SARS-CoV-2 through the postpandemic period.” Science. 2020 May 22;368(6493):860–868. doi: 10.1126/science.abb5793. Epub 2020 Apr 14.

36) Chinazzi M, Davis JT, Ajelli M, Gioannini C, Litvinova M, Merler S, Pastore Y Piontti A, Mu K, Rossi L, Sun K, Viboud C, Xiong X, Yu H, Halloran ME, Longini IM Jr, Vespignani A. (2020) “The effect of travel restrictions on the spread of the 2019 novel coronavirus (COVID-19) outbreak.” Science. 2020 Apr 24;368(6489):395–400. doi: 10.1126/science.aba9757. Epub 2020 Mar 6.

